# Probiotics for the Treatment of Pediatric Allergic Rhinitis: A Systematic Review and Network Meta-Analysis

**DOI:** 10.64898/2025.12.15.25342032

**Authors:** Haiyan Li, Zeyu Chen, Lingyue Guo, Deng Liu, Dongmei Li, Xiaodong Jia, Keqiang Yan

**Author notes:** Corresponding author, Address: Building B, Building 3, Liansheng Technology Park, Binhai High-tech Zone, Tianjin.

## Abstract

**Objective:** This study aimed to evaluate the efficacy, safety, and optimal strains of probiotics for pediatric allergic rhinitis (AR) using meta-analysis and network meta-analysis.

**Methods:** A systematic search was conducted in databases including PubMed, Web of Science, Cochrane Library, and China National Knowledge Infrastructure up to July 31, 2025, to identify randomized controlled trials (RCTs). Inclusion criteria were pediatric patients with AR, probiotic interventions, control groups receiving placebo or standard treatment, and reported outcomes such as Total Nasal Symptom Score (TNSS), Pediatric Rhinoconjunctivitis Quality of Life Questionnaire (PRQLQ), serum IgE levels, clinical efficacy, or adverse events. Study quality was assessed using the JADAD scale, with meta-analysis and network meta-analysis (NMA) performed via RevMan and R software, calculating standardized mean differences (SMD), relative risks (RR), and surface under the cumulative ranking curve (SUCRA) values.

**Results:** Twenty-six RCTs were included, involving 3,014 patients (1,565 in the probiotic group and 1,404 in the control group). Meta-analysis showed that probiotics significantly reduced TNSS (SMD = −0.85, 95% CI [−1.25, −0.44], P < 0.05), improved PRQLQ scores (SMD = −3.94, 95% CI [−4.55, −3.33], P < 0.05), enhanced clinical efficacy (RR = 1.16, 95% CI [1.07, 1.25], P < 0.05), and decreased adverse events (RR = 0.22, 95% CI [0.06, 0.82], P < 0.05), but exerted no overall effect on serum IgE (SMD = −0.39, 95% CI [−0.99, 0.09], P = 0.11). Subgroup and NMA analyses indicated that mixed strains performed superiorly across multiple outcomes.

**Conclusions:** Probiotics, particularly mixed strains, are a safe and effective adjunctive therapy for pediatric AR, improving symptoms and quality of life. Large-scale RCTs are required to validate optimal regimens.

## 1 Introduction

Allergic rhinitis (AR) is a common IgE-mediated inflammatory disorder characterized by nasal itching, congestion, rhinorrhea, and sneezing. Globally, AR affects hundreds of millions of individuals, imposing a substantial economic burden, including medical costs, absenteeism from work or school, and lost productivity. Children are particularly susceptible, with global prevalence rates ranging from 10% to 40% based on epidemiological surveys. The overall physician-diagnosed prevalence is 10.48%, while the self-reported current (within the past 12 months) prevalence stands at 18.12% (1). This elevated prevalence significantly impairs children’s academic performance, quality of life, and psychological well-being, and may progress to complications such as asthma, resulting in long-term respiratory issues (2). Early intervention is essential, as untreated pediatric AR can exacerbate the atopic march and increase the risk of allergic diseases in adulthood (3). Standard treatments include antihistamines, intranasal corticosteroids, and immunotherapy, but these have limitations such as adverse effects, poor adherence, and variable efficacy. For instance, intranasal corticosteroid sprays may induce headaches, nasal mucosal irritation, epistaxis, and cough, whereas oral antihistamines can cause drowsiness and cognitive impairments, particularly in children (4). Prolonged use of these medications may also heighten the risks of drug resistance and systemic side effects, prompting a focus on exploring safe, non-invasive adjunctive therapeutic strategies.

In recent years, the role of the gut microbiome in immune regulation has garnered increasing attention. The gut-nose axis, as an integral component of the immune system, suggests that microbiome dysbiosis may promote Th2-type immune responses, contributing to the onset and progression of AR (5). Probiotics, as live microbial supplements, can alleviate allergic reactions by strengthening the intestinal barrier, balancing Th1/Th2 responses, and promoting regulatory T cell (Treg) activity (6). Several randomized controlled trials (RCTs) have shown that strains such as Lactobacillus spp. and Bifidobacterium spp. reduce AR symptoms, though results vary: some report improvements, while others find no effects (7). Previous meta-analyses have predominantly focused on adults or mixed populations and have not adequately addressed strain specificity (8). Systematic evidence pertaining to pediatric AR remains limited, with a notable absence of network meta-analysis (NMA) to compare the relative efficacy of different strains (9). This study employs systematic literature retrieval and meta-analysis to evaluate the efficacy, safety, and superior strains of probiotics in pediatric AR, with the aim of providing evidentiary support for clinical practice.

## 2 Materials and Methods

### 2.1 Literature Search Strategy

A computerized literature search was conducted to identify RCTs investigating probiotic interventions for pediatric allergic rhinitis, spanning from database inception to July 31, 2025. Databases searched encompassed Chinese resources including China National Knowledge Infrastructure (CNKI), Wanfang Database, China Science and Technology Journal Database, China Biology Medicine disc (CBMdisc), and National Science and Technology Library (NSTL) and international platforms incorporating Web of Science, PubMed, Embase, and Cochrane Library. We searched these databases using subject and free words: (((“Rhinitis, Allergic”) OR (((allergic rhinitis) OR (Allergic Rhinitides)) OR (Allergic Rhinitis))) AND ((“Child”) OR (Children))) AND ((“Probiotics”) OR (Probiotic)). The language limits set for studies were those published in English or Chinese. Additional studies were identified through manual screening of reference lists from included articles to ensure comprehensive coverage.

### 2.2 Literature inclusion and exclusion criteria

Inclusion criteria: 1) Study design: RCTs investigating probiotic interventions for AR, including parallel or crossover designs, regardless of blinding status, published in Chinese or English. 2) Participants: Pediatric patients diagnosed with AR. 3) Intervention: Probiotic formulations (e.g., Lactobacillus, Bifidobacterium, Saccharomyces, or Mixed-strain) administered via any route (oral, nasal, etc.), with no restrictions on dosage, administration route, or treatment duration. 4) Control groups: Placebo with identical dosage, standard therapy (e.g., antihistamines, intranasal corticosteroids), or no intervention. 5) Outcomes: At least one primary or secondary outcome reported. Primary outcomes included Total Nasal Symptom Score (TNSS), Pediatric Rhinoconjunctivitis Quality of Life Questionnaire (PRQLQ), and adverse outcomes. Secondary outcomes encompassed immunological parameters, such as serum IgE levels and eosinophil counts before and after treatment.

Exclusion criteria: 1) Non-RCT studies, including case reports, case series, or retrospective studies. 2) Studies lacking a control group or unreported outcome data. 3) Studies focusing on non-allergic rhinitis (e.g., infectious rhinitis) or non-nasal allergic conditions. 4) Conference abstracts or studies with inaccessible full texts.

### 2.3 Selection of studies and data extraction

Following duplicate removal using EndNote 20 software, two independent researchers rigorously screened and extracted data in accordance with predefined inclusion and exclusion criteria. The results were cross-verified, with discrepancies resolved through third-party adjudication. Extracted data were recorded in a Excel spreadsheet, encompassing the following domains: 1) Study characteristics: First author, publication year, and country of origin. 2) Study design: Trial type (e.g., parallel, crossover) and blinding methodology (if applicable). 3) Participant details: Sample size and age range of enrolled subjects. 4) Intervention parameters: Probiotic strain(s), dosage regimen, and treatment duration. 5) Outcomes.

### 2.4 Quality assessment of the included studies

The methodological rigor of included studies was evaluated using the Jadad Scale (10), which assesses the following domains: 1) Random sequence generation. 2) Allocation concealment. 3) Blinding procedures. 4) Documentation of withdrawals and dropouts. Each criterion was scored independently, with the cumulative score representing the overall quality assessment outcome. Studies achieving a Jadad score ≥3 were classified as high-quality literature. Additionally, the Cochrane Risk of Bias Tool (11) was employed by two independent researchers to evaluate the risk of bias across seven domains: random sequence generation, allocation concealment, blinding of participants and personnel, blinding of outcome assessment, incomplete outcome data, selective reporting, and other potential biases. Each domain was rated as low, unclear or high risk of bias, and the overall risk of bias for individual studies was determined through a synthesis of domain-specific evaluations.

### 2.5 Statistical analysis

Meta-analyses were performed using Review Manager 5.3.0 (RevMan) and R 4.5.1 software. Heterogeneity was assessed via the I^2^ statistic, with I^2^ >50% indicating substantial heterogeneity and warranting a random-effects model, while I^2^ ≤ 50% supported the use of a fixed-effects model. Subgroup analyses stratified by probiotic strains (e.g., Lactobacillus, Bifidobacterium) were conducted to explore potential sources of heterogeneity. NMA was further implemented to compare probiotics efficacy across strains, with ranking probabilities quantified using the Surface Under the Cumulative Ranking Curve (SUCRA). The SCURA values ranges from 0 to 100%, with larger values indicating a higher ranking of the strain among all strains (12). Statistical significance was defined as P <0.05.

## 3 Results

### 3.1 Literature screening results

The initial search yielded 1,351 articles. After removing duplicates and screening titles and abstracts, 98 articles were retained. Full-text review and application of inclusion/exclusion criteria resulted in the final inclusion of 26 studies (Figure 1), comprising 6 Chinese-language and 20 English-language publications. All studies were RCTs, collectively involving 3,014 participants: 1,565 participants in probiotics intervention groups and 1,404 in control groups (placebo or standard therapy). Key characteristics of the included studies are summarized in Table 1.

**Figure 1.**
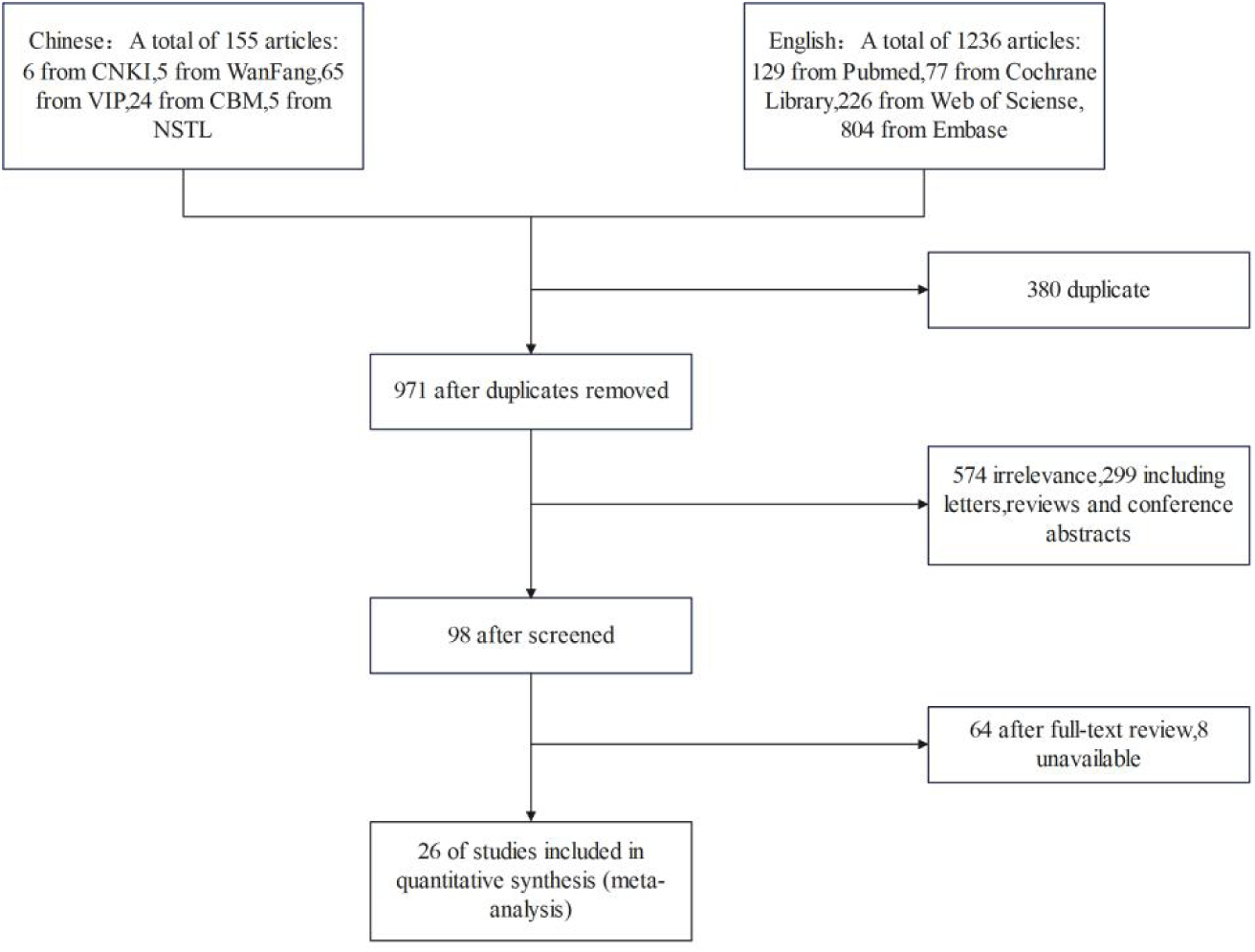
Flowchart of the search process for article included.

**Table 1.**
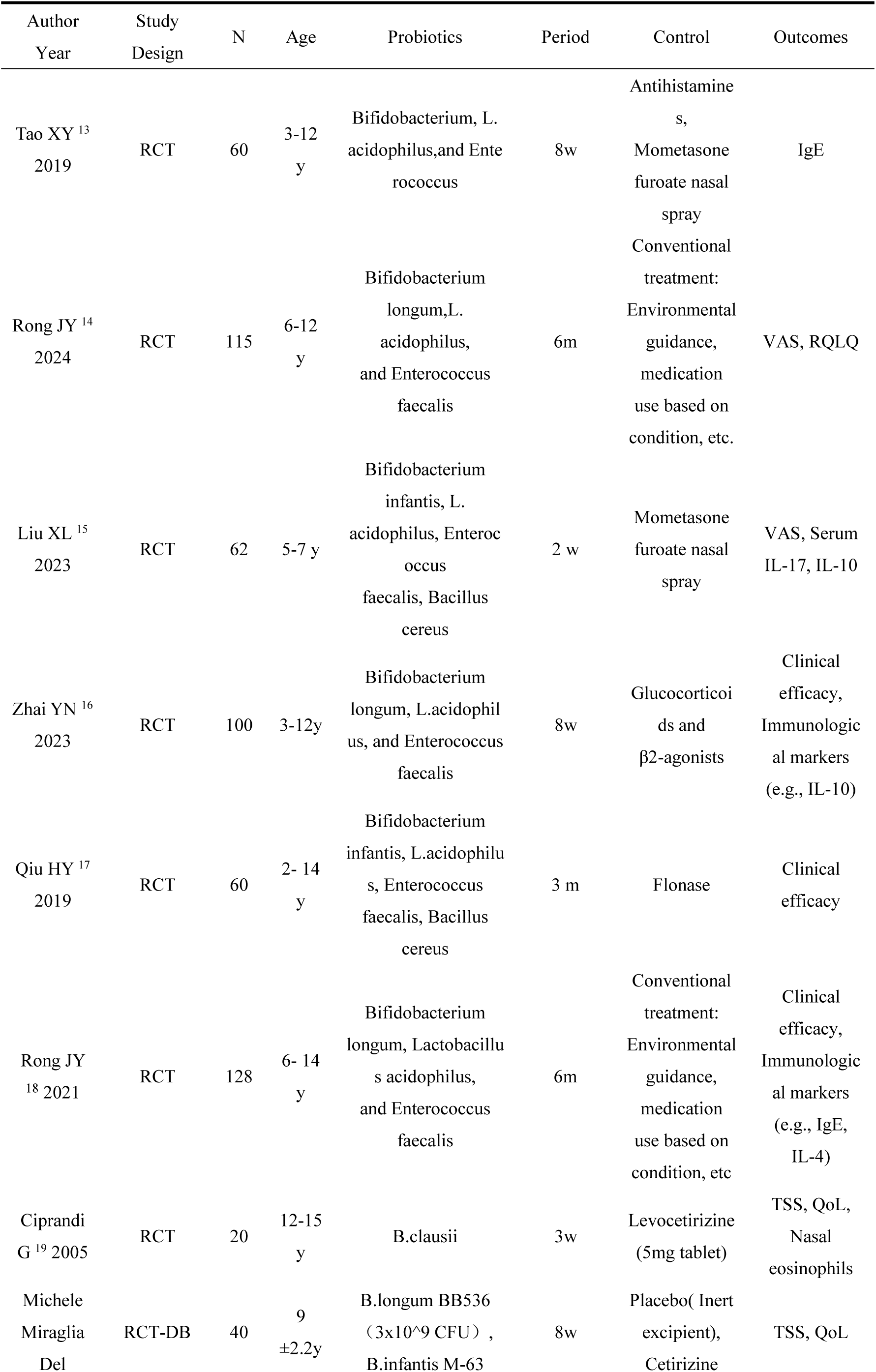
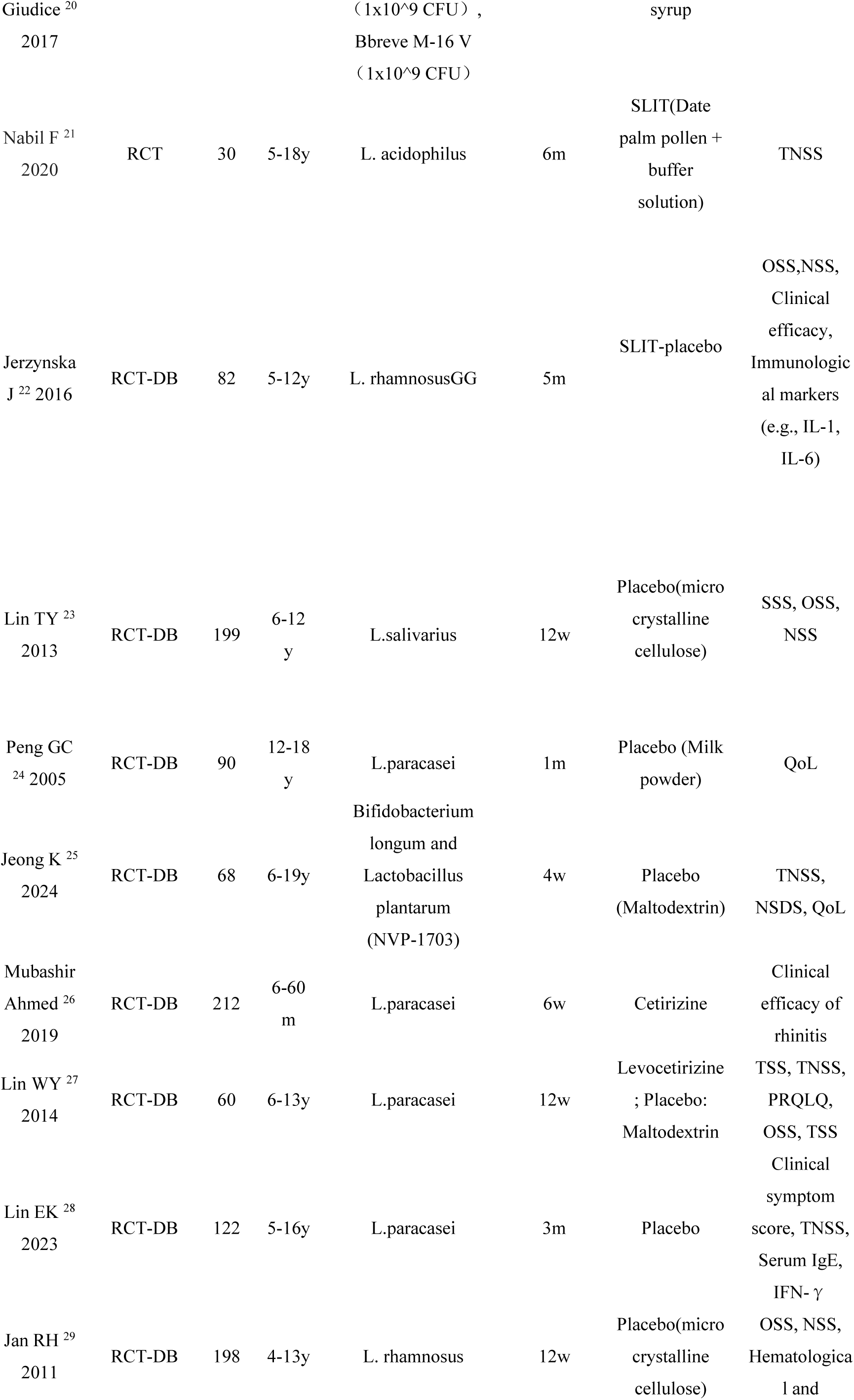
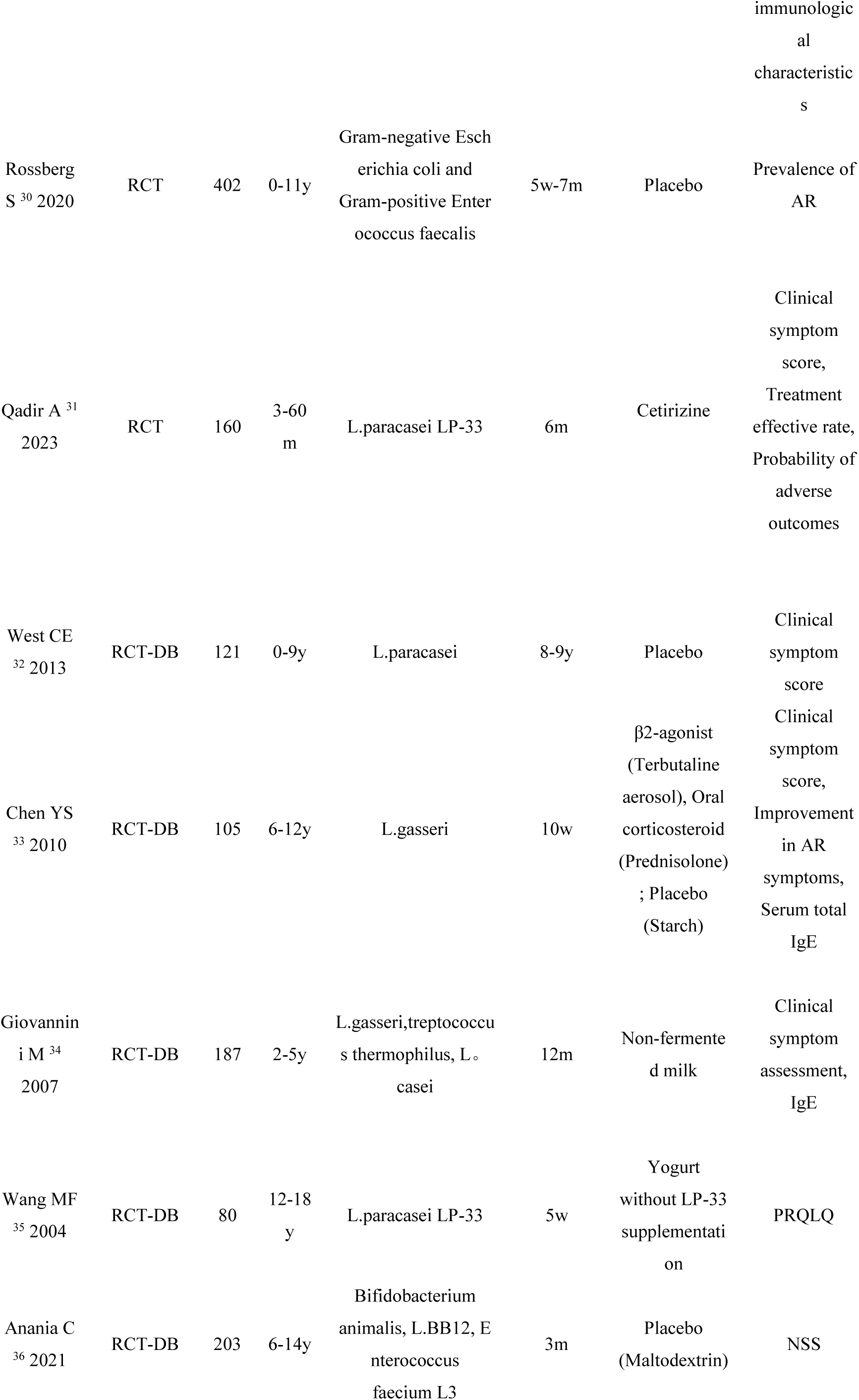
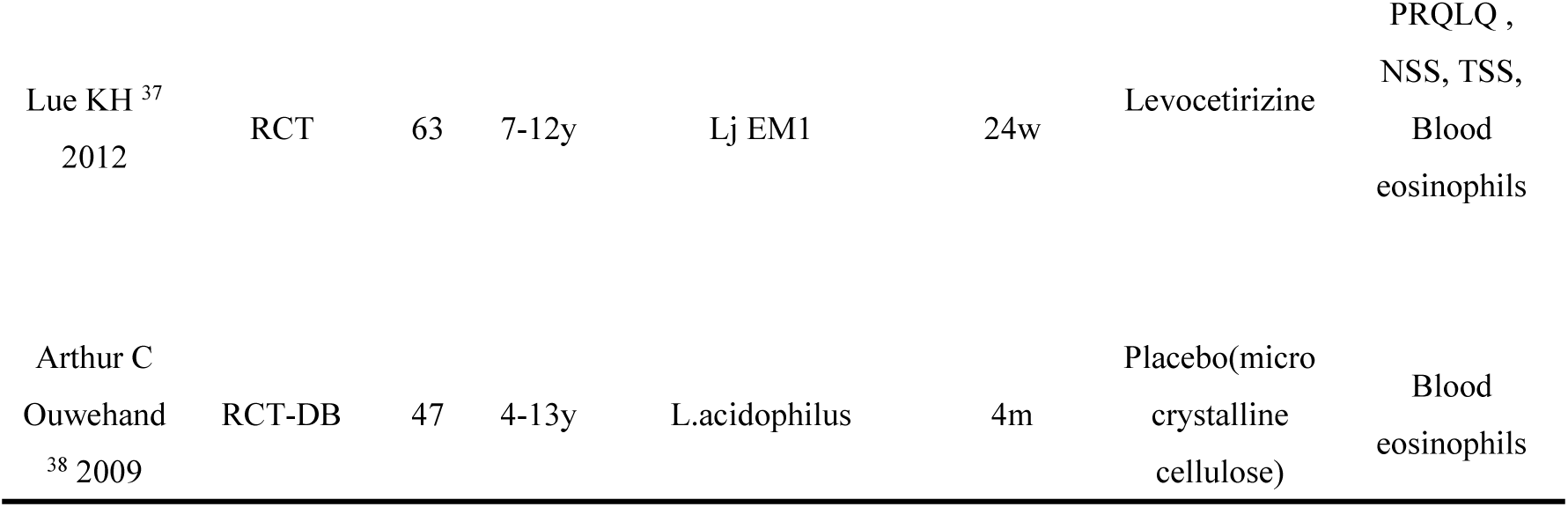
Baseline characteristics of the studies included.

### 3.2 Quality assessment

Methodological quality evaluation using the Jadad scale identified 20 studies as high-quality literature (Table 2). Risk of bias assessment conducted via the Cochrane Risk of Bias Tool revealed 16 studies with low risk, 1 studies with unclear risk, and 9 studies with high risk of bias (summarized in Figure 2 and detailed in Figure 3).

**Figure 2.**
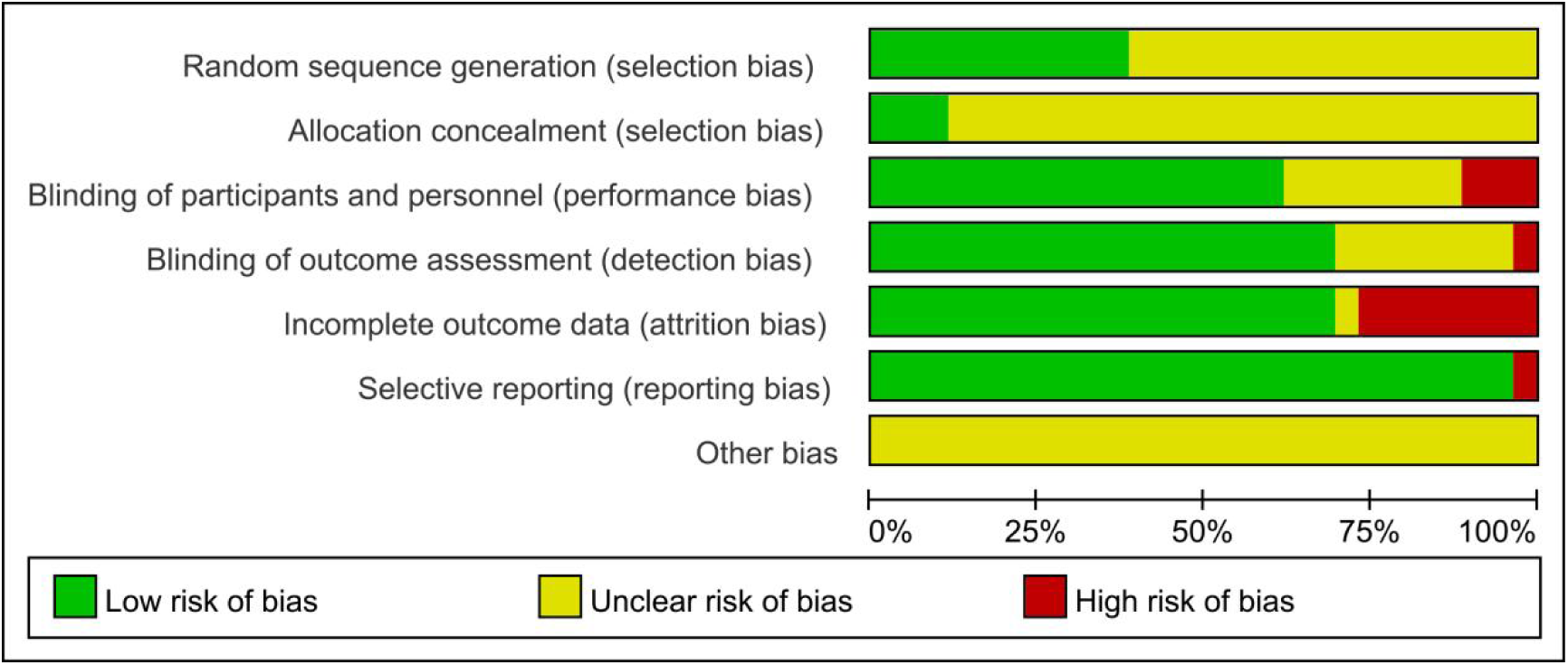
Risk of bias summary of included studies.

**Figure 3.**
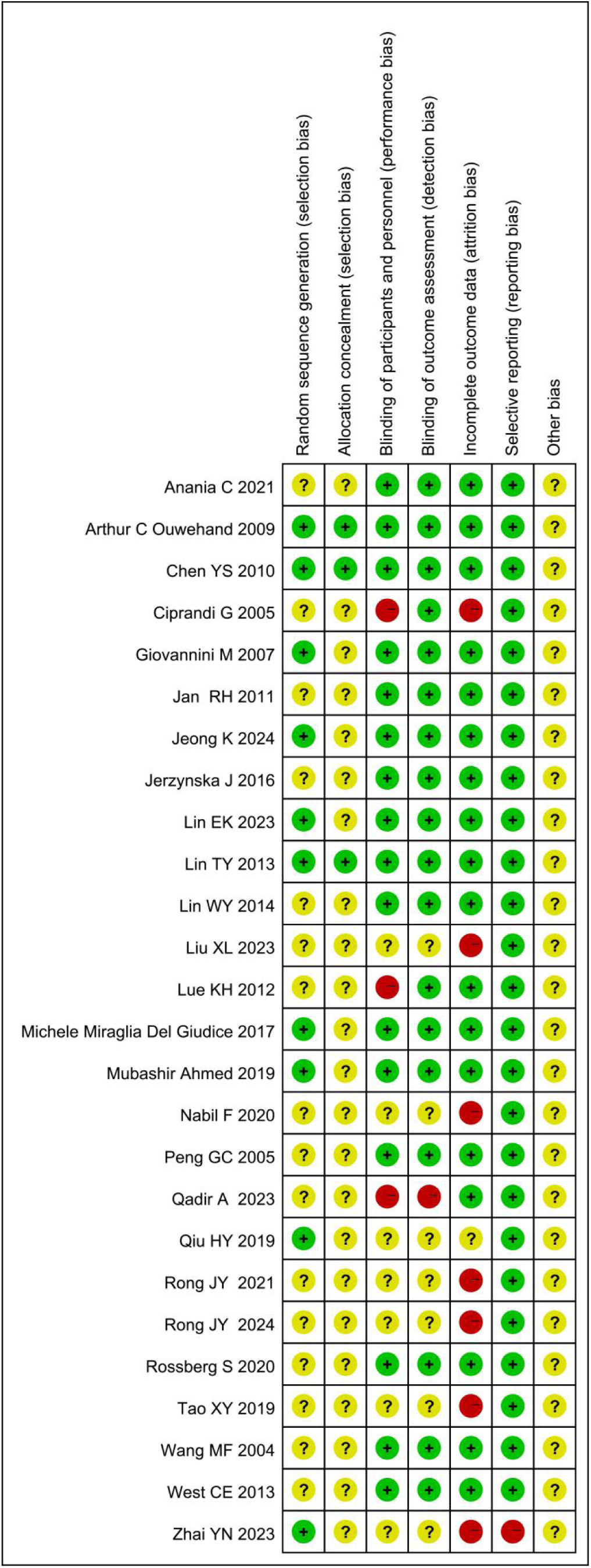
Risk of bias assessment for included studies.

**Table 2.**
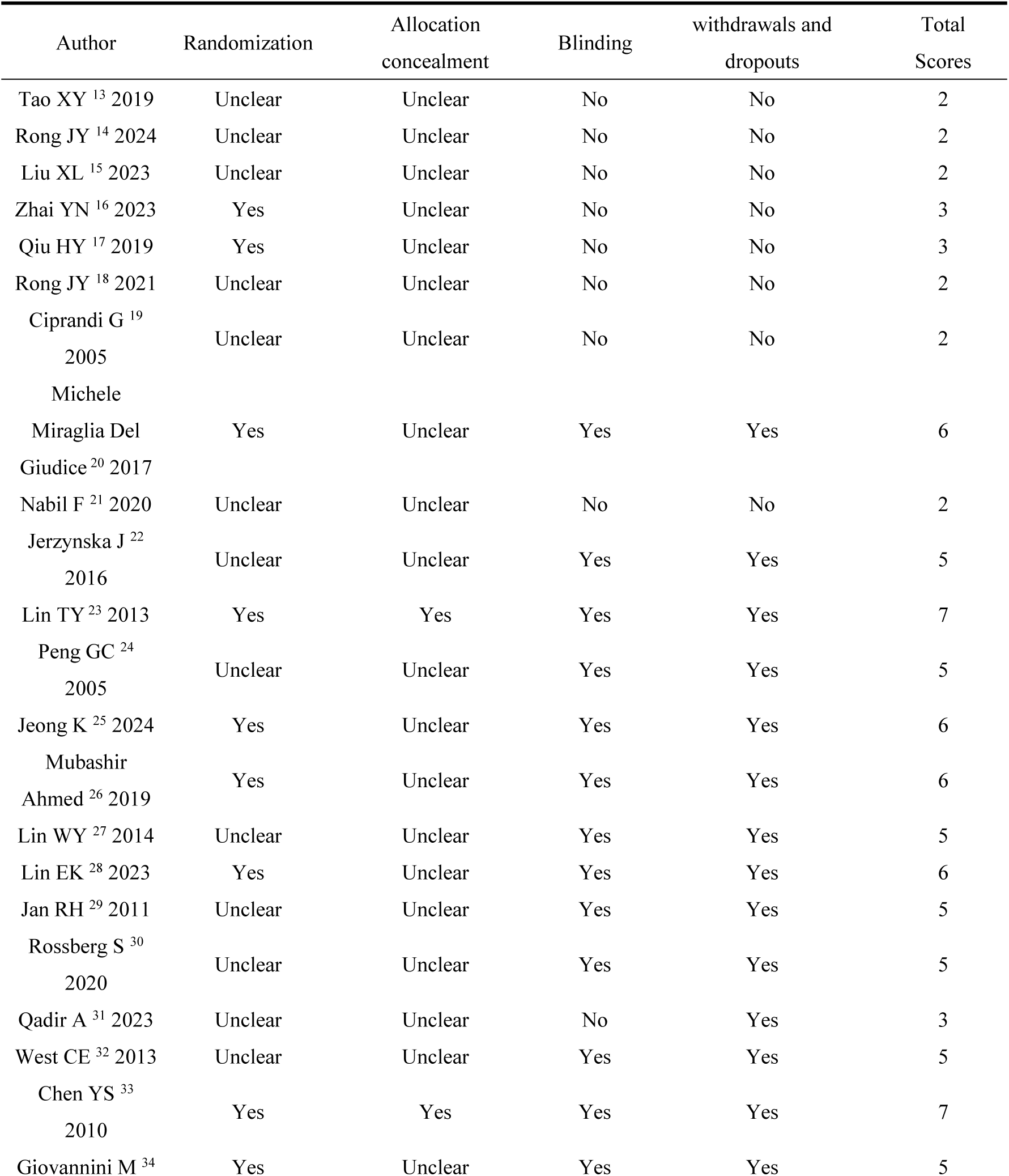
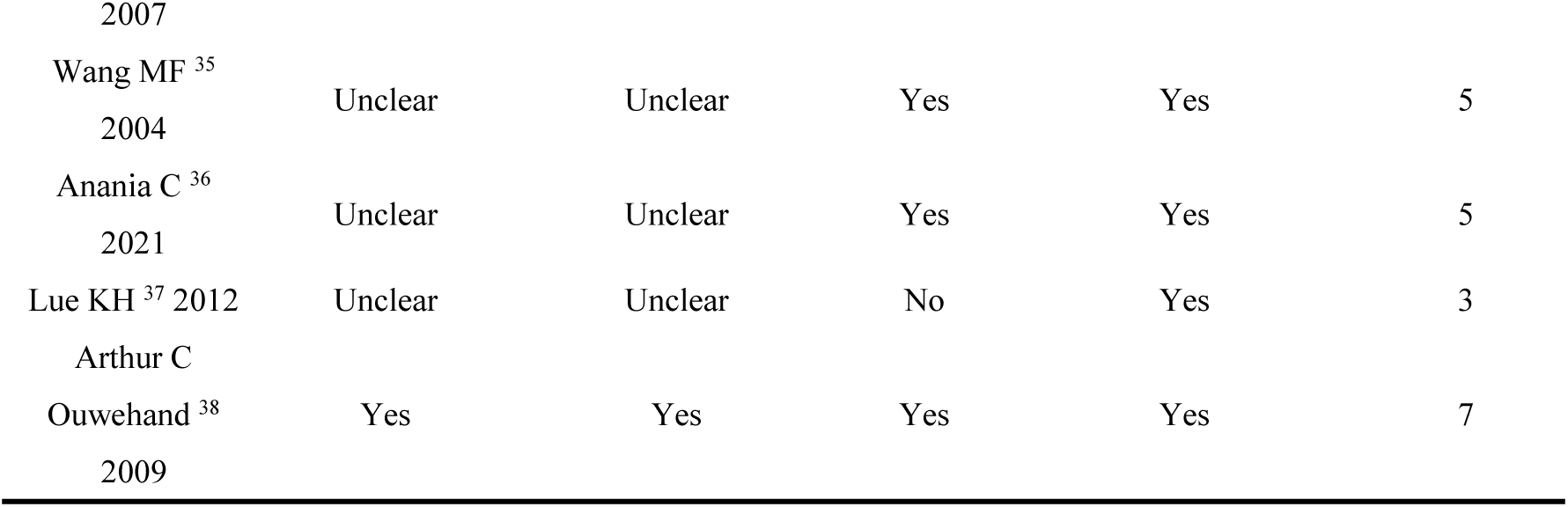
Jadad scale of the including studies.

### 3.3 Results of meta-analysis

#### 3.3.1 Effects of probiotics on TNSS

Four studies (18,21,23,25) reported post-treatment TNSS comparisons between probiotics and control groups. Heterogeneity testing revealed significant between-study variation (I^2^ =72%, P=0.01), prompting adoption of a random-effects model. The pooled analysis demonstrated statistically significant reductions in TNSS scores favoring probiotic interventions (standardized mean difference [SMD]=-0.85, 95%CI [−1.25, −0.44], P<0.0001), as illustrated in Figure 4.

**Figure 4.**
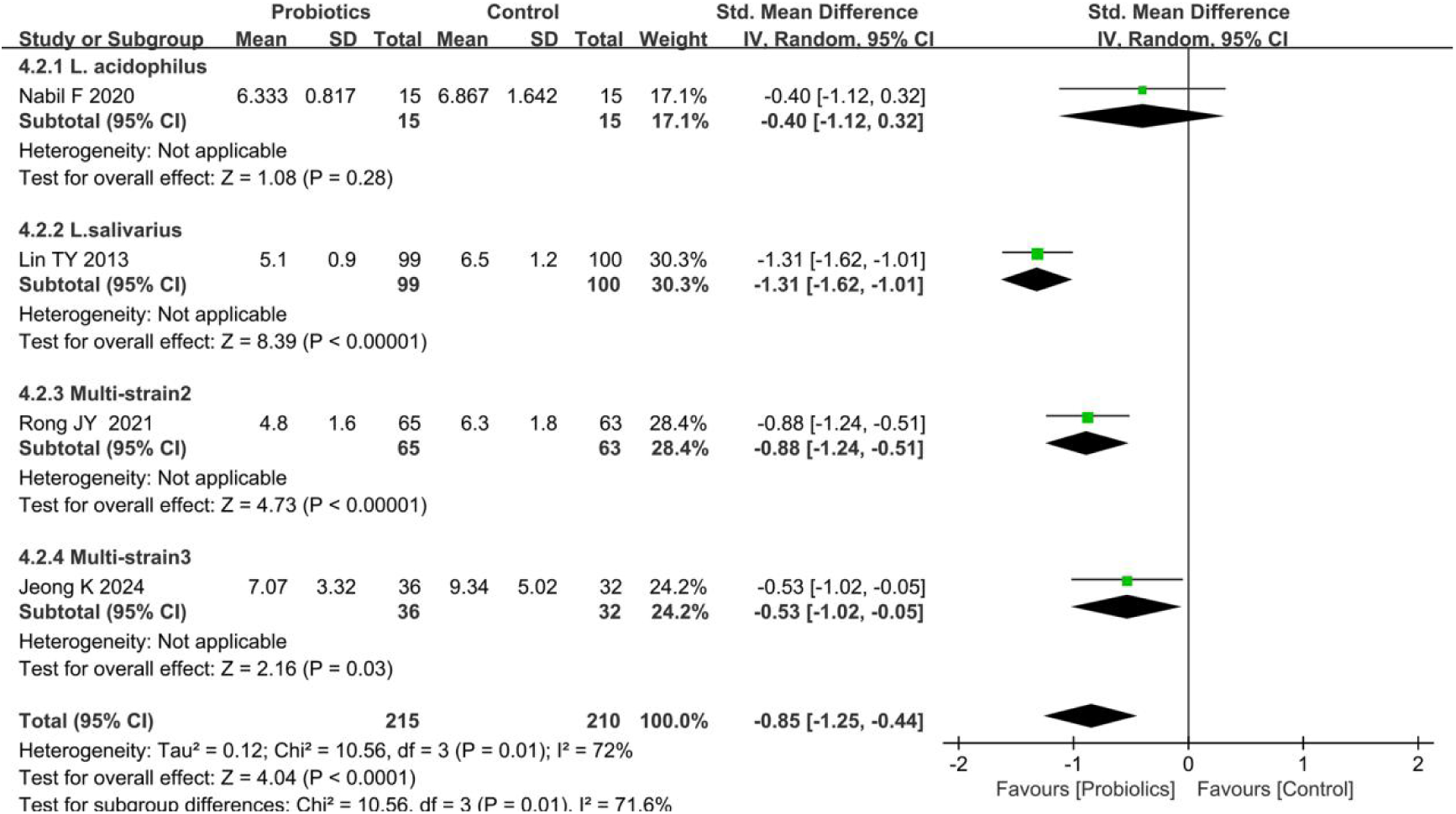
Forest plot comparing the effect of probiotics and control on TNSS. Subgroup by the types of probiotics.

Subgroup analyses were conducted according to the usage of probiotics. Two studies (13,25) evaluating multi-strain formulations (Multi-strain2: B. longum, L. acidophilus, and E. faecalis; Multi-strain3: B. longum and L. plantarum [NVP-1703]) both reported statistically significant TNSS difference between probiotics and controls (P<0.05). In contrast, two studies (21,23) assessing single-strain interventions showed divergent outcomes: L. acidophilus exhibited no significant TNSS improvement (P=0.28), whereas L. salivarius demonstrated clinically meaningful reductions (P<0.05; Figure 4).

#### 3.3.2 Effects of probiotics on serum IgE

Seven studies (13,16,18,25,28,33,34) reported serum IgE levels following probiotics interventions compared to controls. Heterogeneity testing indicated substantial between-study variation (I^2^=89%, P<0.0001), necessitating a random-effects model. Pooled analysis revealed no statistically significant difference in post-treatment serum IgE level between probiotics and control groups (SMD=-0.39, 95% CI [−0.99, −0.09], P=0.11), as shown in Figure 5.

**Figure 5.**
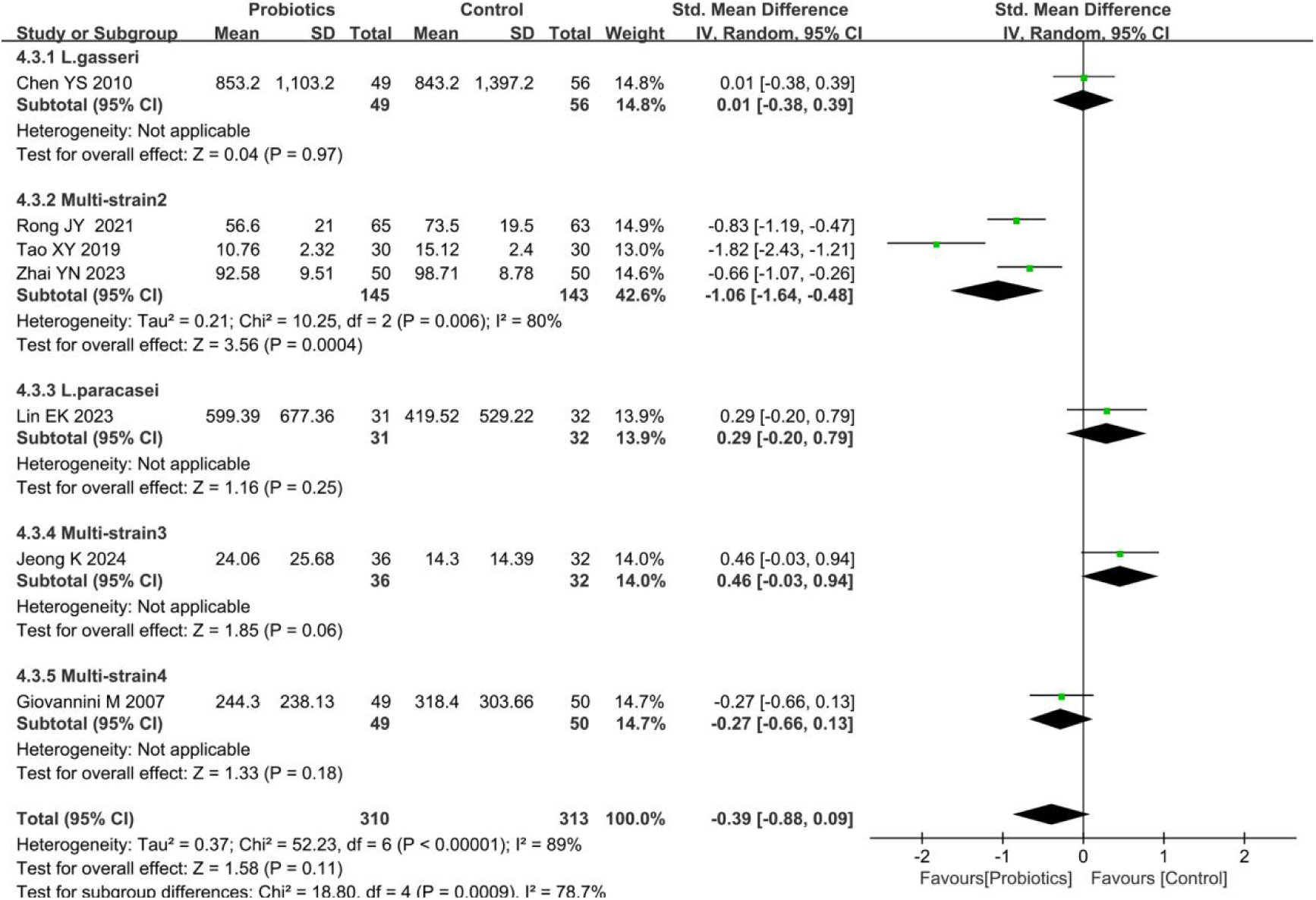
Forest plot comparing the effect of probiotics and control on the serum IgE. Subgroup by the types of probiotics.

Subgroup analyses stratified by probiotic strains demonstrated heterogeneous effects. Three studies (13,16,18) evaluating Multi-strain2 (B. longum, L. acidophilus, and E. faecalis) exhibited significant heterogeneity (I^2^=80%, P<0.01), with a random-effects model showing statistically significant difference in serum IgE levels between two groups (SMD=-1.06, 95% CI [−1.64, −0.48], P<0.001). In contrast, two studies (25,34) assessing Multi-strain3 (B. longum and L. plantarum [NVP-1703]) and Multi-strain4 (L. gasseri, S. thermophilus, and L. casei) found no significant differences in IgE levels compared to controls (P=0.06 and P=0.18, respectively). Similarly, two additional studies28,33 investigating L. gasseri and L. paracasei interventions reported no statistically significant IgE modulation (P=0.97 and P=0.25, respectively; Figure 5).

#### 3.3.3 Effects of probiotics on PRQLQ

Two studies (24,35) reported post-treatment PRQLQ scores between probiotics and control groups. Heterogeneity testing revealed significant between-study variation (I^2^=85%, P=0.010), warranting a random-effects model. The pooled analysis demonstrated a statistically significant improvement in PRQLQ scores favoring probiotic interventions (SMD=-3.94, 95% CI [−4.55, −3.33], P<0.05), as illustrated in Figure 6.

**Figure 6.**
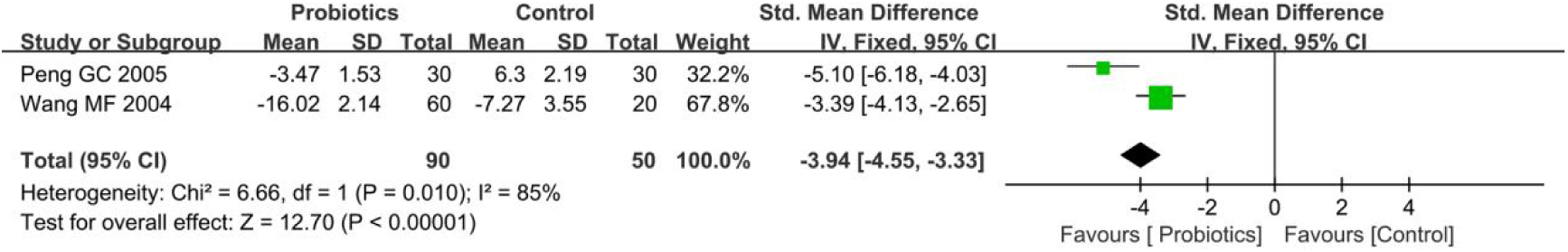
Forest plot comparing the effect of probiotics and control on PRQLQ.

#### 3.3.4 Effects of probiotics on clinical efficacy

Six studies (16–18,26,31,33) evaluated the therapeutic efficacy of probiotic interventions compared to controls. Heterogeneity testing revealed no significant between-study variation (I^2^=19%, P=0.29), supporting the use of a fixed-effects model. Pooled analysis demonstrated a statistically significant improvement in clinical efficacy favoring probiotic groups (risk ratio [RR]=1.16, 95% CI [1.07, 1.25], P<0.05), as detailed in Figure 7.

**Figure 7.**
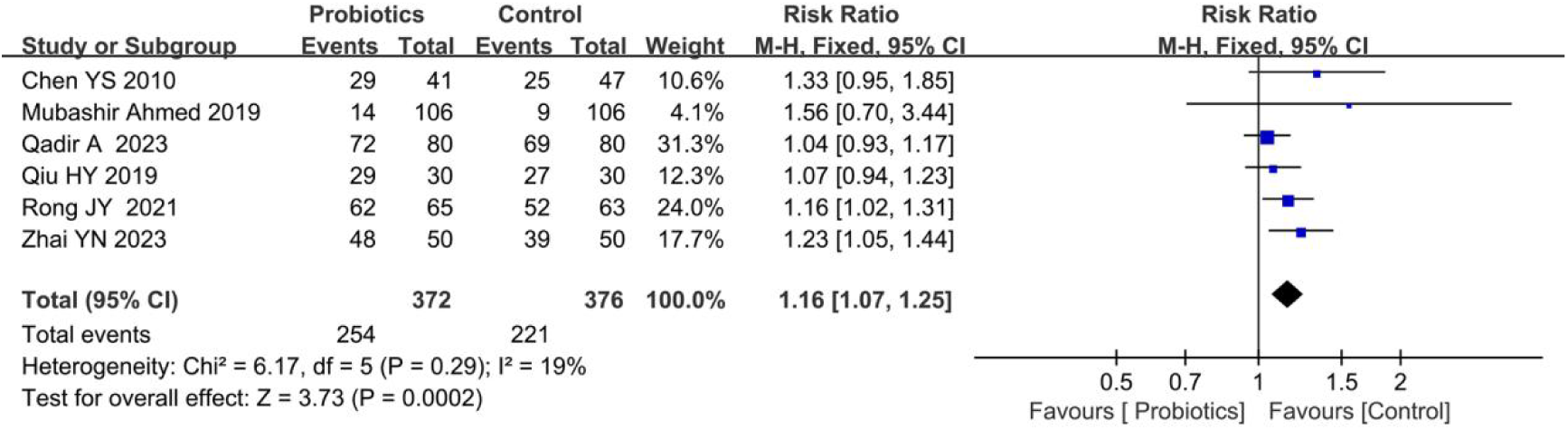
Forest plot comparing the effect of probiotics and control on clinical efficacy.

#### 3.3.5 Effects of probiotics on adverse outcomes

Adverse outcomes, including headache, nausea, epistaxis, vomiting, pharyngitis, and abdominal pain during treatment, were reported in two studies (16,31) comparing probiotic and control groups. Heterogeneity testing indicated no significant between-study variation (I^2^=0%, P=0.95), justifying the use of a fixed-effects model. Pooled analysis revealed a statistically significant differences in adverse outcomes between two groups (RR=0.22, 95% CI [0.06, 0.82], P<0.05), as illustrated in Figure 8.

**Figure 8.**
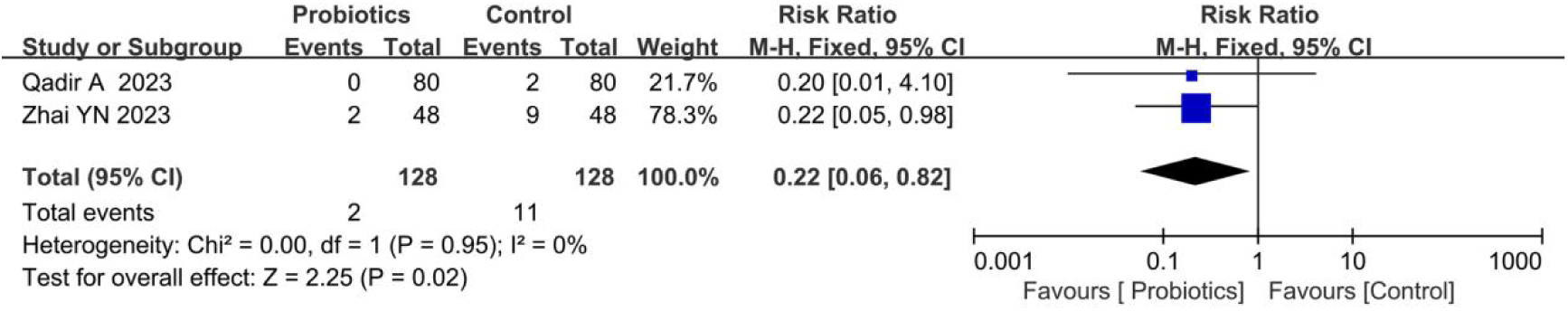
Forest plot comparing the effect of probiotics and control on adverse outcomes.

### 3.4 SCURA analysis identified superior probiotic strains

In the NMA, network connectivity was ensured by requiring all interventions to be indirectly comparable through a shared comparator. To achieve this, all non-probiotic interventions were consolidated into a unified “Control” group within the NMA framework. This approach was predicated on the assumption that all studies were conducted under similar standard-of-care protocols, with probiotics serving as the sole variable. The consolidation aimed to evaluate the relative efficacy of distinct probiotic strains compared to conventional management strategies.

#### 3.4.1 Rank of diverse probiotic strains on TSS

SUCRA analysis revealed that Multi-strain1 (B. longum subsp. longum BB536, B. infantis M-63, and B. breve M-16) and Bacillus clausii demonstrated superior efficacy in reducing TSS compared to conventional therapy, with ranking probabilities as follows: Multi-strain1 (SUCRA=100.0%) > B. clausii (SUCRA=40.5%) > conventional therapy (SUCRA=9.5%) (Figure 9).

**Figure 9.**
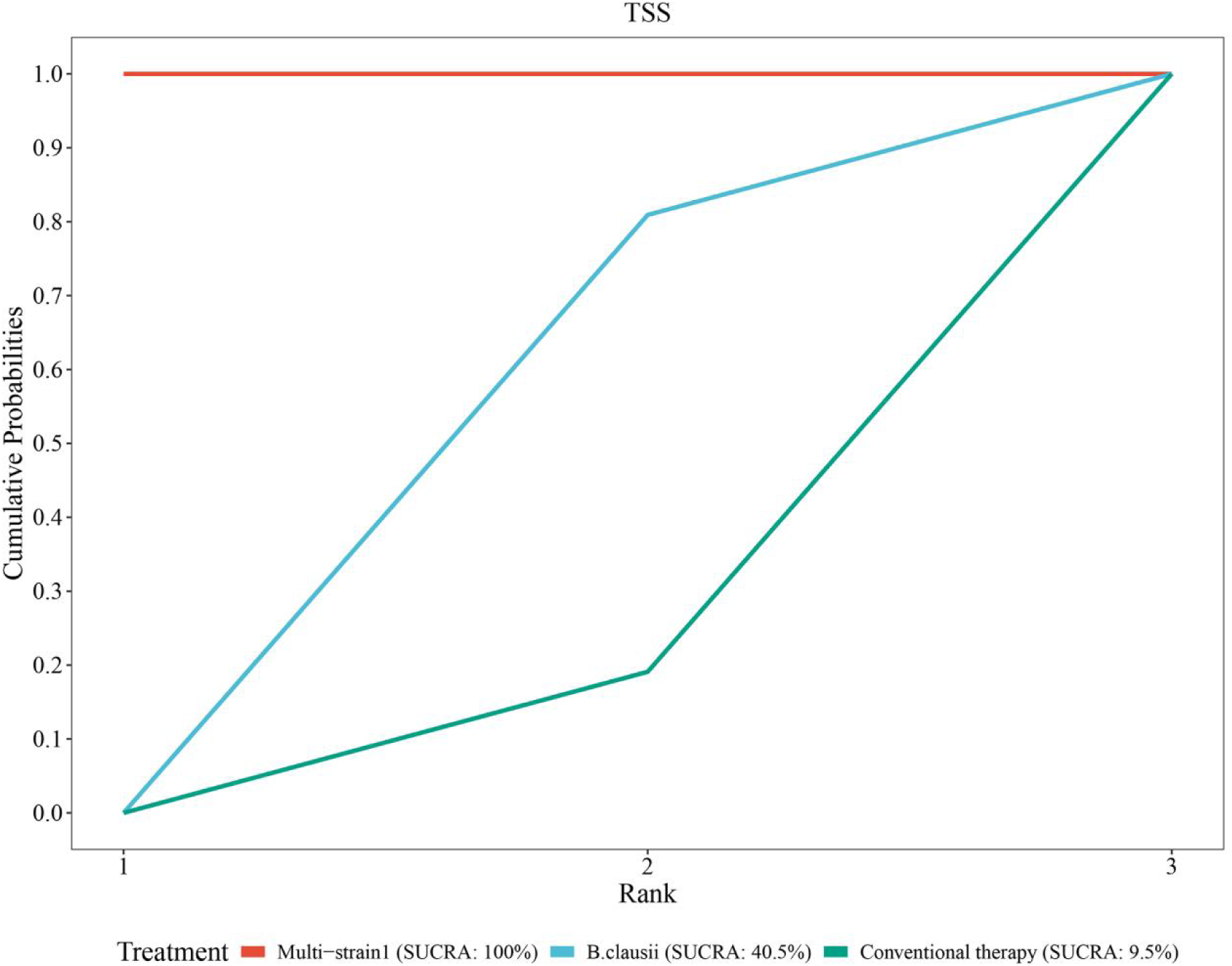
SCURA probability rank of TSS.

#### 3.4.2 Rank of diverse probiotic strains on clinical efficacy

SUCRA analysis demonstrated superior efficacy of multi-strain probiotics and specific Lactobacillus strains over conventional therapy. The ranking probabilities were as follows: Multi-strain2 (B. longum, L. acidophilus, and E. faecalis; SUCRA=88.4%) > Multi-strain5 (B. infantis, L. acidophilus, E. faecalis, and B. cereus; SUCRA=63.6%) > L. gasseri (SUCRA=53.2%) > L. paracasei (SUCRA=37.2%) > conventional therapy (SUCRA=7.6%) (Figure 10).

**Figure 10.**
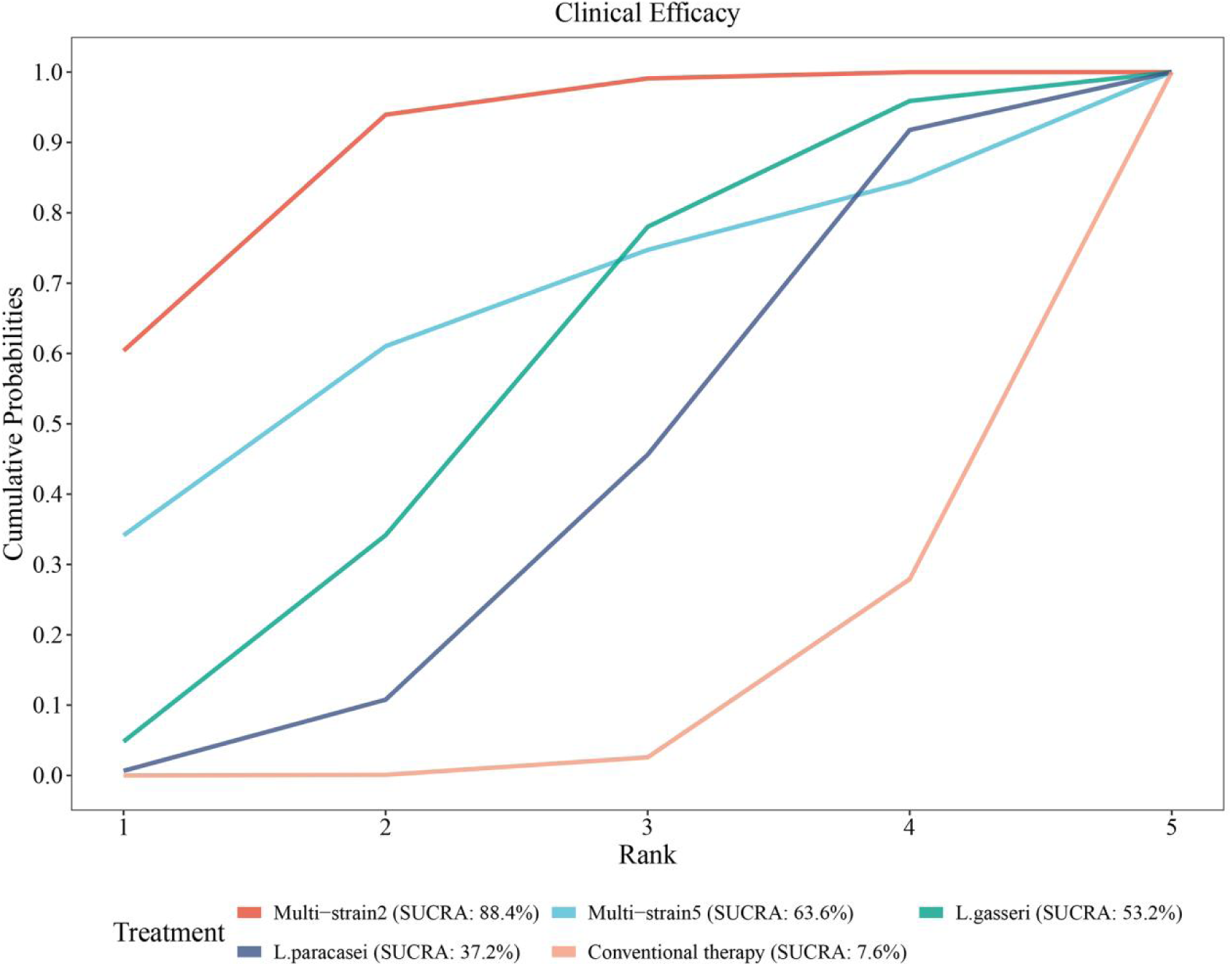
SCURA probability rank of clinical efficacy.

#### 3.4.3 Rank of diverse probiotic strains on adverse outcomes

SUCRA analysis disclosed superior efficacy of Multi-strain2 and L. paracasei compared to conventional therapy. The ranking probabilities were as follows: Multi-strain2 (B. longum, L. acidophilus, and E. faecalis; SUCRA=77.0%) > L. paracasei (SUCR=62.5%) > conventional therapy (SUCRA=10.4%) (Figure 11).

**Figure 11.**
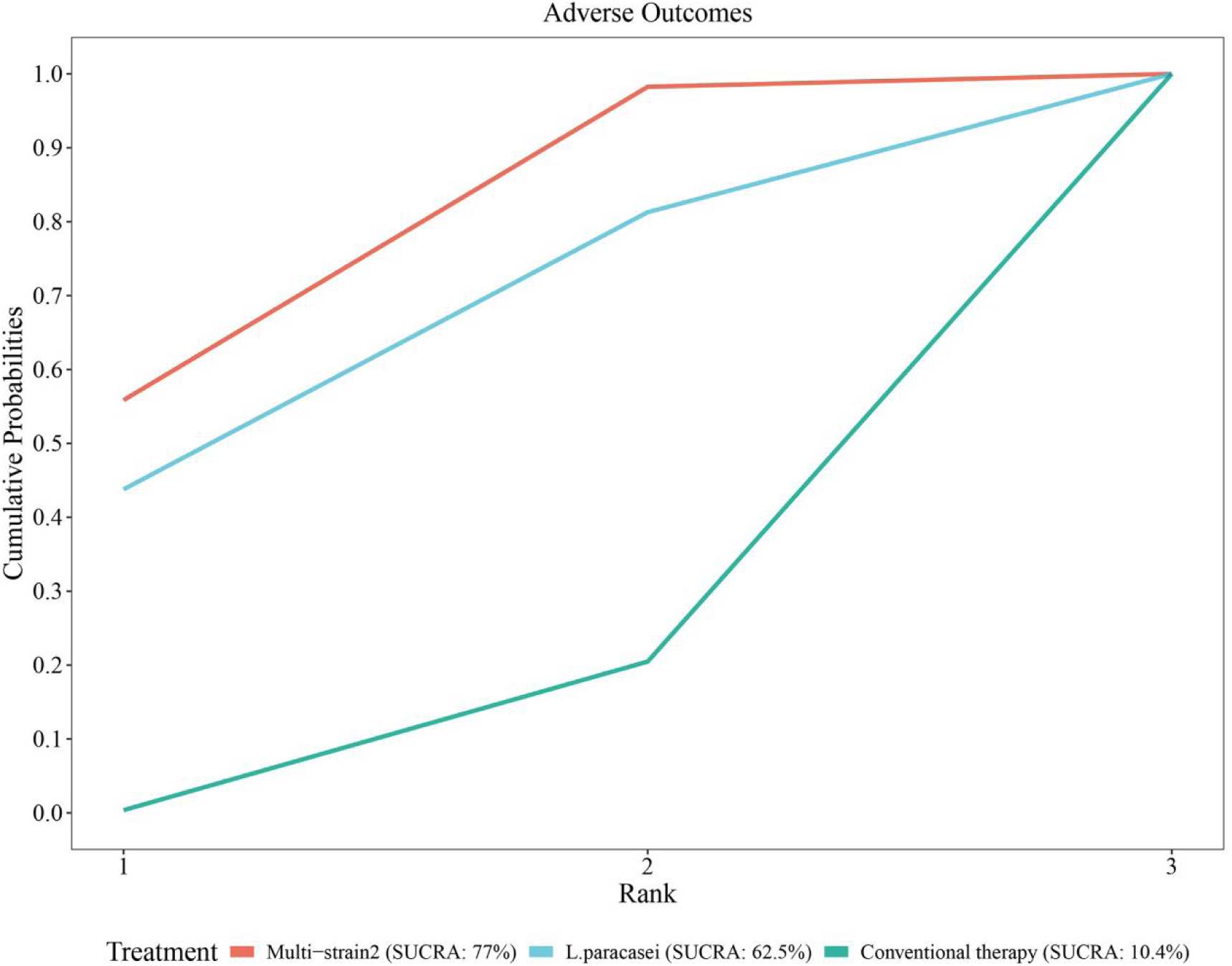
SCURA probability rank of adverse outcomes.

#### 3.4.4 Rank of diverse probiotic strains on TNSS

SUCRA analysis exposed superior efficacy of multi-strain probiotics and Lactobacillus strains in reducing TNSS compared to conventional therapy. The ranking probabilities were as follows: Multi-strain3 (B. longum and L. plantarum [NVP-1703]; SUCRA=86.8%) > Multi-strain4 (L. gasseri, S. thermophilus, and L. casei; SUCR=70.5%) > L. salivarius (SUCRA=63.6%) > L. acidophilus (SUCRA=25.5%) > conventional therapy (SUCRA=3.6%) (Figure 12).

**Figure 12.**
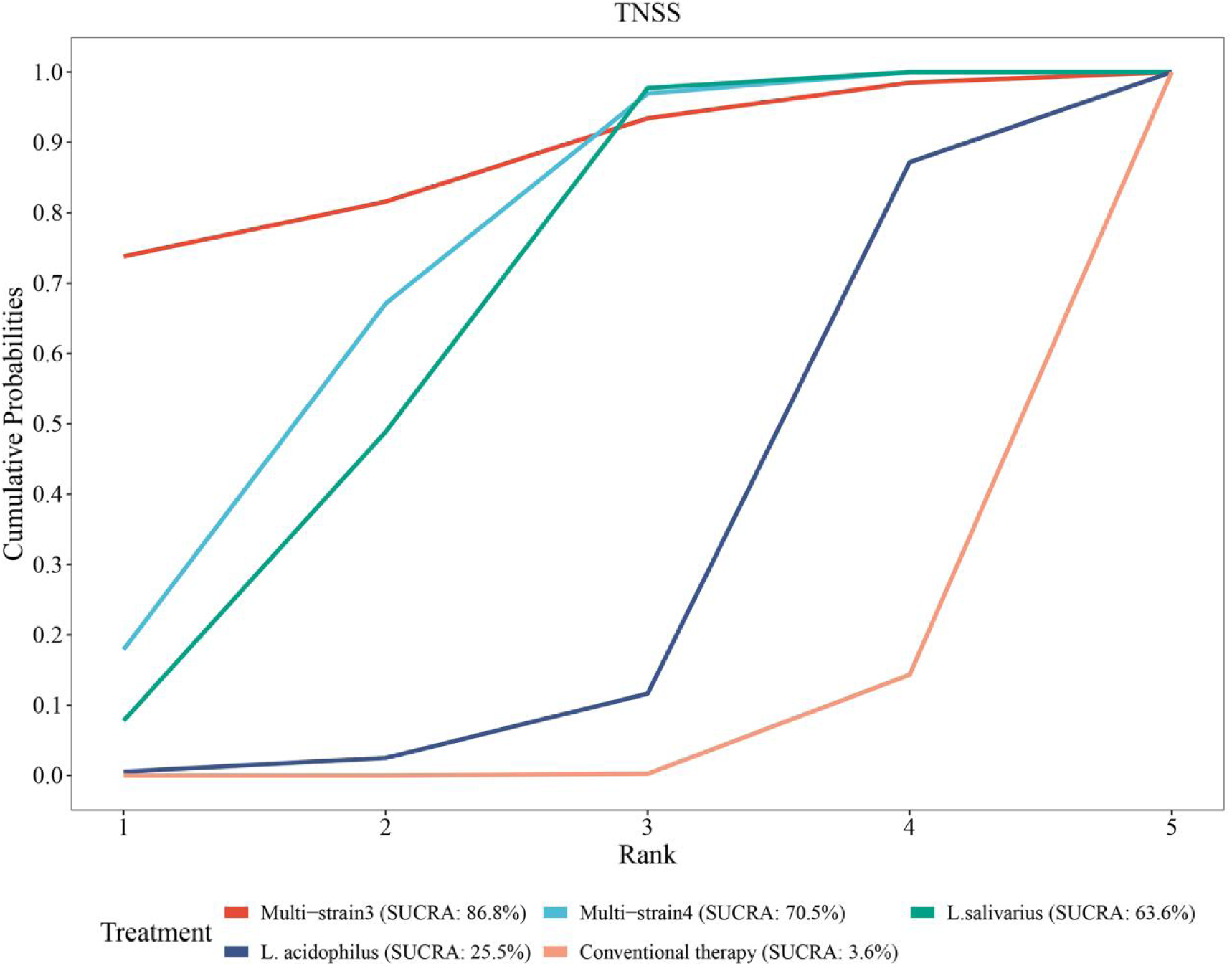
SCURA probability rank of TNSS.

#### 3.4.5 Rank of diverse probiotic strains on serum IgE

SUCRA analysis revealed superior efficacy of multi-strain probiotics and L. gasseri in reducing serum IgE levels compared to conventional therapy, while conventional therapy outperformed Multi-strain3 and L. paracasei. The ranking probabilities were as follows: Multi-strain4 (L. gasseri, S. thermophilus, and L. casei; SUCRA=92.1%) > Multi-strain2 (B. longum, L. acidophilus, and E. faecalis; SUCRA=74.6%) > L. gasseri (SUCRA=51.4%) > conventional therapy (SUCRA=49.32%) > Multi-strain3 (B. longum and L. plantarum [NVP-1703]; SUCRA=21.4%) > L. paracasei (SUCRA=11.2%) (Figure 13).

**Figure 13.**
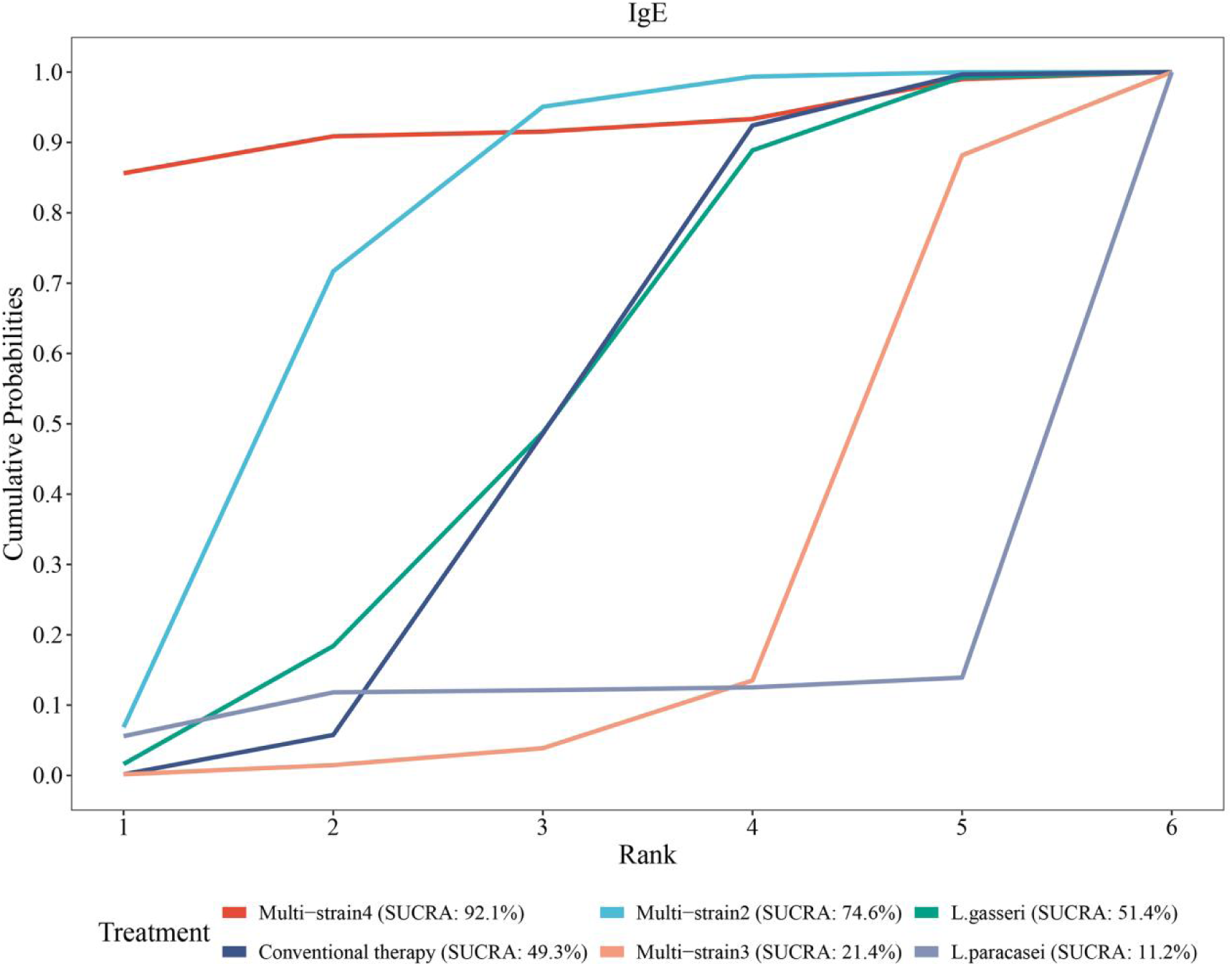
SCURA probability rank of the serum IgE.

## 4 Discussion

This meta-analysis incorporated 26 randomized controlled trials, encompassing a total of 3,014 pediatric patients with AR, to systematically evaluate the efficacy and safety of probiotic therapy. The findings indicate that probiotic treatment significantly reduces nasal symptom scores, enhances rhinitis-related quality of life, improves clinical efficacy, and decreases the incidence of adverse reactions, although it exerts no overall significant impact on serum total IgE levels. These results provide evidentiary support for probiotics as an adjunctive therapy for pediatric allergic rhinitis, aligning with existing literature while underscoring the importance of strain specificity and heterogeneity.

First, regarding the amelioration of nasal symptoms, this study revealed that the TNSS in the probiotic group was significantly lower than in the control group, consistent with outcomes from several prior meta-analyses. For instance, Zajac et al. (39) reported in a meta-analysis of adult and pediatric allergic rhinitis that probiotics effectively alleviate nasal symptoms, such as nasal congestion, itching, and sneezing. Subgroup analyses further demonstrated that mixed strains outperformed single strains in reducing TNSS, potentially attributable to synergistic effects that enhance immune modulation and intestinal barrier function. SUCRA analysis confirmed that Multi-strain3 (SUCRA=86.8%) and Multi-strain4 (SUCRA=70.2%) ranked highest in TNSS improvement, suggesting a preference for mixed probiotic formulations in clinical practice. However, single strains such as L. acidophilus exhibited non-significant effects (P=0.28), which may relate to strain specificity, dosage, and treatment duration, necessitating further optimization.

Second, with respect to immunological parameters, this analysis showed no overall significant reduction in serum total IgE levels with probiotics, although certain mixed strains demonstrated notable differences in subgroup analyses. SUCRA analysis indicated that Multi-strain4 (SUCRA=92.1%) performed best in reducing IgE, whereas L. paracasei ranked poorly (SUCRA=11.2%), emphasizing the critical role of strain selection. Probiotics may exert their effects through regulation of the Th1/Th2 balance, promotion of regulatory T cell (Treg) activity, and reduction of inflammatory cytokines; however, their modulation of IgE is not universally applicable and should be evaluated in conjunction with individual immune status.

In terms of quality of life and clinical efficacy, this study confirmed that probiotics significantly improve the total PRQLQ score and clinical outcomes, echoing the systematic review by Güvenç et al. (40), which demonstrated enhanced quality of life in patients with allergic rhinitis. SUCRA analysis highlighted the superiority of Multi-strain2 (SUCRA=88.4%) in clinical efficacy, suggesting that mixed strains may indirectly alleviate rhinitis symptoms by improving gut microbiome diversity. Furthermore, the incidence of adverse reactions was significantly lower in the probiotic group, with common events such as abdominal pain or headache occurring less frequently, supporting the safety of probiotics and aligning with the meta-analysis by Wang et al. (41), which reported lower rates of probiotic-related adverse events compared to conventional medications.

Despite the positive findings of this study, several limitations persist. First, inter-study heterogeneity was substantial, with I^2^ values of 72% for TNSS and 89% for IgE, potentially arising from strain diversity, dosage variations, inconsistent treatment durations, and baseline patient characteristics. Although random-effects models and subgroup analyses mitigated this issue, future research should standardize intervention protocols. Second, most studies were short-term, lacking long-term follow-up data, thereby precluding assessment of the sustained effects of probiotics. Additionally, the samples were primarily derived from Asian and European populations, with an absence of data from African or Latin American regions, limiting the generalizability of the results. Finally, while SUCRA analysis facilitates strain ranking, it relies on indirect comparisons, and the efficacy of treatments requires further investigation.

In conclusion, this study demonstrates that probiotics, particularly mixed strains, represent a safe and effective adjunctive therapeutic option for pediatric allergic rhinitis, significantly improving symptoms and quality of life, while their impact on immunological parameters is strain-dependent. Future research should prioritize large-scale, multicenter, long-term studies to explore optimal strain combinations, dosages, and mechanisms, incorporating diverse populations to further validate these findings and inform clinical practice.

## Author Contributions

HY Li conceptualised this systematic review. ZY Chen and LY Guo searched the literature and screened the titles and abstracts. KQ Yan and XD Jia reviewed all full-text articles and extraced data. HY Li and D Liu formally analysed, visualised and validated the data. KQ Yan supervised the systematic review. All authors were responsible for the methodology and review and editing of the manuscript. All authors had final responsibility for the decision to submit the manuscript for publication.

## Data Availability Statement

Data sharing is not applicable to this article as no datasets were generated or analyzed during this study.

